# Face masks, old age, and obesity explain country’s COVID-19 death rates

**DOI:** 10.1101/2020.06.22.20137745

**Authors:** D. Miyazawa

## Abstract

We tested the hypothesis that not wearing face masks, old age, and obesity can largely explain COVID-19 death rates across countries. In the regression analysis, they contributed to 61.5%, 40.5%, and 43.8%, respectively, and a model including all these variables contributed to 70.2% of the variation in the cumulative number of COVID-19 deaths per million on May 13, 2020, in 22 countries. We also proposed the hypothesis that these variables may be confounders of other suspected factors with large differences between Western and non-Western countries, such as the BCG vaccination policy. These results contribute to the elucidation of differences in COVID-19 death rates between countries and support a suspected causal effect of face masks.

## Main Text

There have been considerable differences in the number of deaths caused by the coronavirus disease (COVID-19) across countries. Western countries, in particular, have recorded a higher number of deaths compared to non-Western countries. Several regression studies have identified explanatory variables for the differences in the number of deaths such as age, obesity, and the previous Bacille Calmette-Guérin (BCG) vaccination^1,2,3^. However, face mask-wearing rates have not been used in cross-country regression studies as an explanatory variable, although face masks have received increasing attention as an effective means to prevent the transmission of COVID-19^4,5,6^. In this study, we tested the hypothesis that face mask-wearing rates can largely explain COVID-19 death rates by country, along with old age and obesity, which are the independent risk factors for hospitalized COVID-19 patients^7,8,9,10^.

We were able to obtain synchronized mask-wearing rates from 22 countries. The number of explanatory variables should generally not exceed much more than 10% of the sample size in a multiple regression analysis^11^; therefore, only the current BCG vaccination policy^12^, and the rates of avoiding crowded public spaces were added as additional explanatory variables. Crowded public spaces are another major factor that are suspected to contribute to differences in COVID-19 death rates by country. Although there was a possibility of unmeasured confounding, it was not this study’s intention to fully prove the causality or independence of the variables, but rather to provide clues to reveal the reason for the difference in COVID-19 death rates by country to the extent that analysis with several variables selected on a medical basis can be revealed. In addition because we noticed a trend in these variables, with clear differences between Western and non-Western countries a priori, we explored possible confounding between these variables.

We first calculated Spearman’s correlations between the variables and the cumulative number of deaths per million since January 1, starting on March 20 (Fig.1A, Data S1) as well as weekly sums of deaths per million starting on March 16 (Fig. 1B, Data S1) to determine variables to include in the regression. High correlations were observed with mask non-wearing rates in March and with no current BCG. The mask non-wearing rate (April–May) showed a moderate correlation. Not avoiding crowds (March) generally showed a low correlation. Not avoiding crowds (April–May) showed a time-dependent increase in correlation with cumulative death, showing a moderate correlation. Age-related variables were highly correlated, consistent with a previous report^8^. The correlation coefficients were lower in the younger age groups (age 65–69 years) than in the higher age groups in both sexes. Body mass index (BMI) showed moderate correlation with a clear sex-dependent difference. Male BMI showed higher correlations than female BMI, corroborating evidence that males are more susceptible to severe outcomes from COVID-19 than women^13,14^. In addition, females have been reported to engage in more preventive behaviors than males^15,16^.

**Fig. 1A and 1B.**
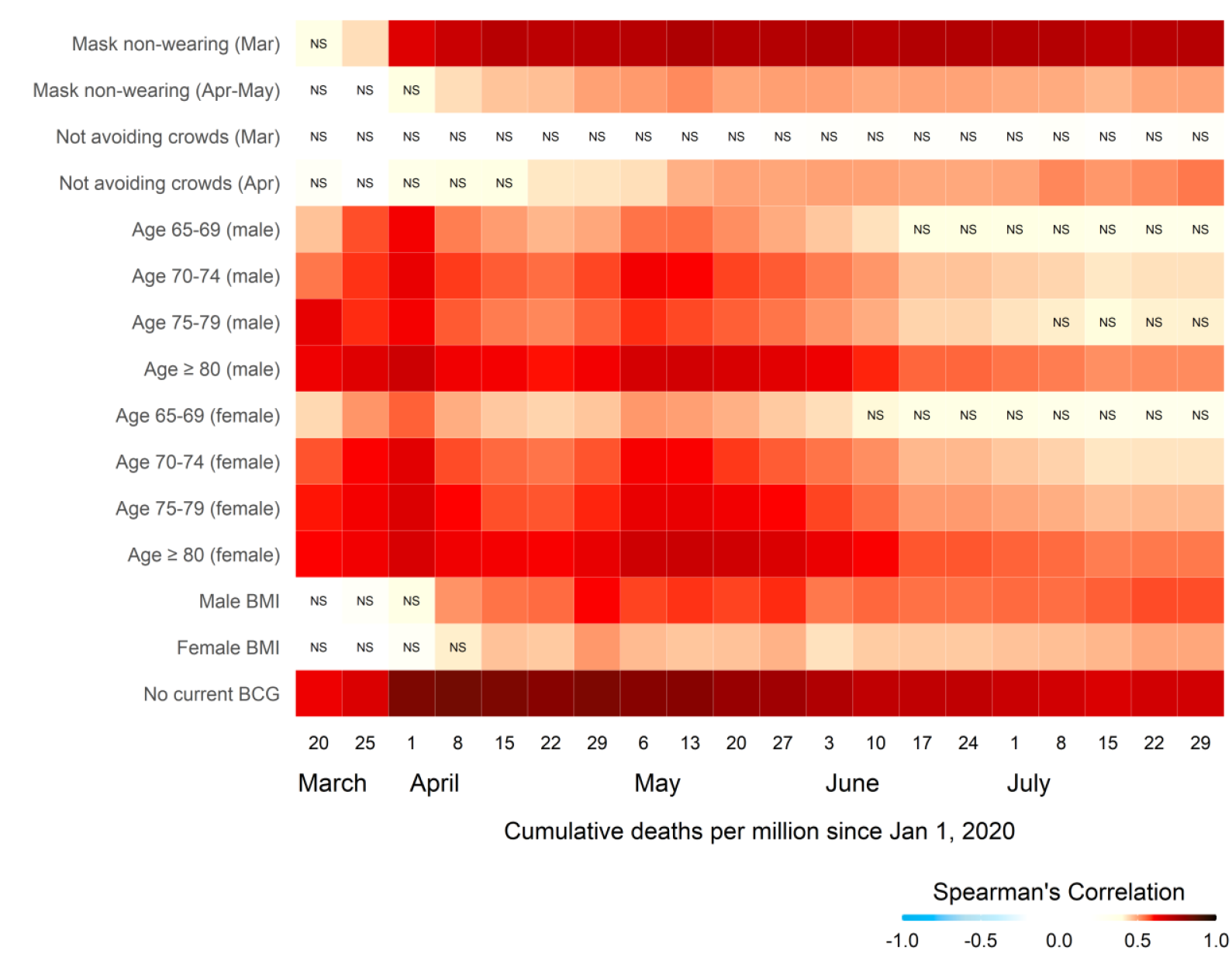
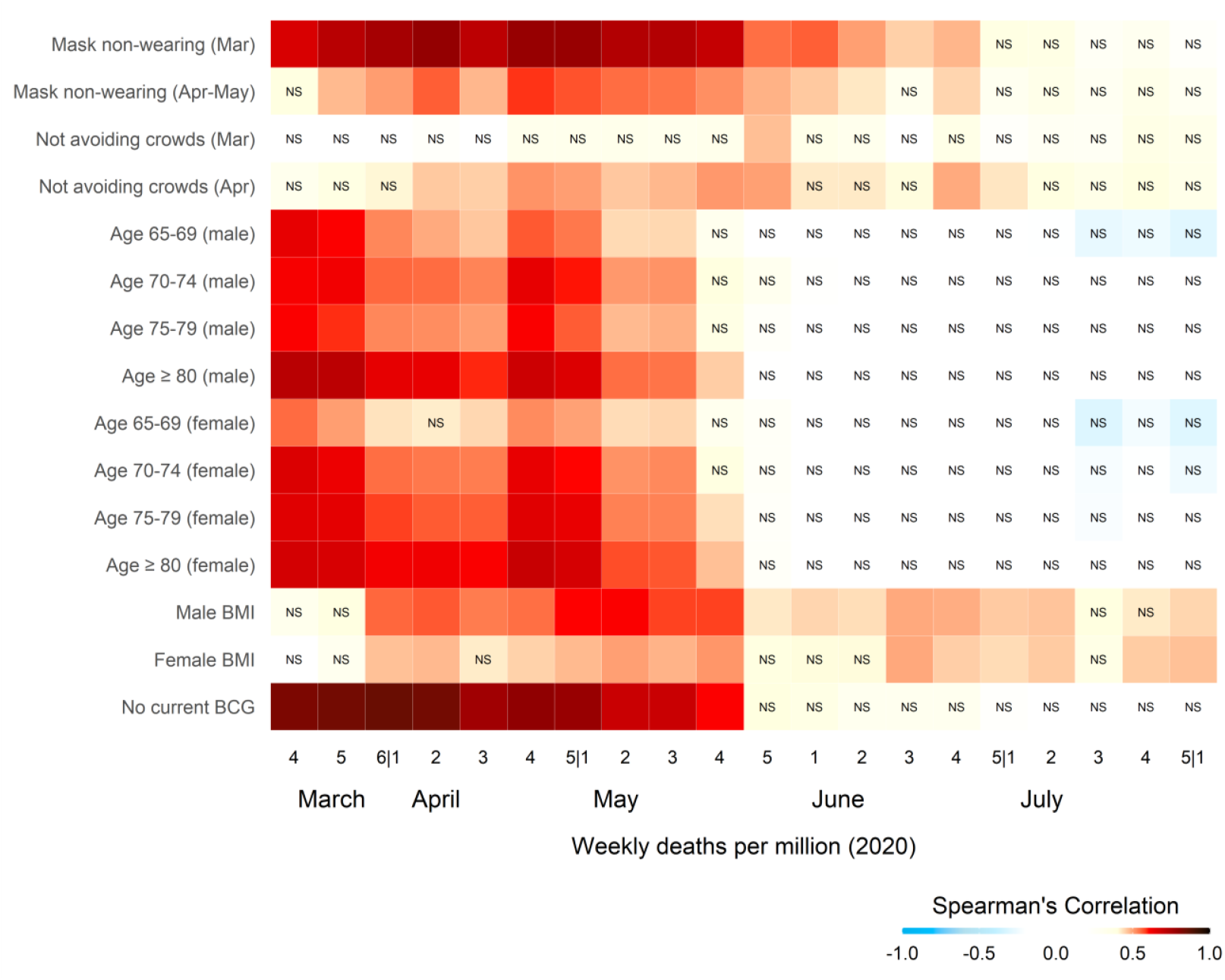
Spearman’s correlation with the cumulative number of deaths (a) Number of weekly deaths (b) Mask-wearing rates in March and April–May were calculated from survey responses during March 9–18, 2020, and April 26–May 1, 2020, respectively. Age-related predictors indicate the percentage of the population in a specified range. BMI, body mass index; NS, not significant (*P* > 0.05).

The 22 countries were divided into 12 Western and 10 non-Western countries, and correlations were examined similarly. Spearman’s correlations between the variables and weekly deaths per million were not significant, in both Western and non-Western countries, possibly because of the small number of countries. Therefore, only the correlations between the cumulative number of deaths are shown. We defined Western countries in this study as North America, Europe, Australia, and New Zealand. None of the 12 Western countries (UK, France, Italy, USA, Spain, Germany, Canada, Sweden, Norway, Finland, Denmark, and Australia) currently offer the BCG vaccine, and all 10 non-Western countries (Mexico, Malaysia, China, Saudi Arabia, India, Indonesia, Philippines, Japan, Singapore, and Thailand) currently do. Therefore, “no current BCG” is equal to “Western” in this study, so the correlation with “no current BCG” was not calculated in this section. In fact, globally, among 24 countries in the world that do not currently provide universal BCG vaccination (Australia, Austria, Belgium, Canada, Czechia, Denmark, Ecuador, Finland, France, Germany, Greece, Israel, Italy, Luxembourg, The Netherlands, New Zealand, Norway, Slovak Republic, Slovenia, Spain, Sweden, Switzerland, and the United States), only Ecuador and Israel are not “Western.”

In Western countries (Fig. 2A, Data S1), only age≥80 years in both sexes showed a significant positive correlation. Interestingly, in Western countries, there was a significant negative correlation in mask non-wearing (April–May). This is attributed to the marked reduction in the mask non-wearing rate in some countries with a high number of deaths, possibly caused by a fear of getting the disease. Possible reasons for the correlations of mask non-wearing rates being low and not significant within Western countries may be due to the small variation between countries and the small number of countries. The standard deviation for the mask non-wearing rate (March) of Western countries was 7.06, compared to 19.2 for non-Western countries.

**Fig. 2A and 2B.**
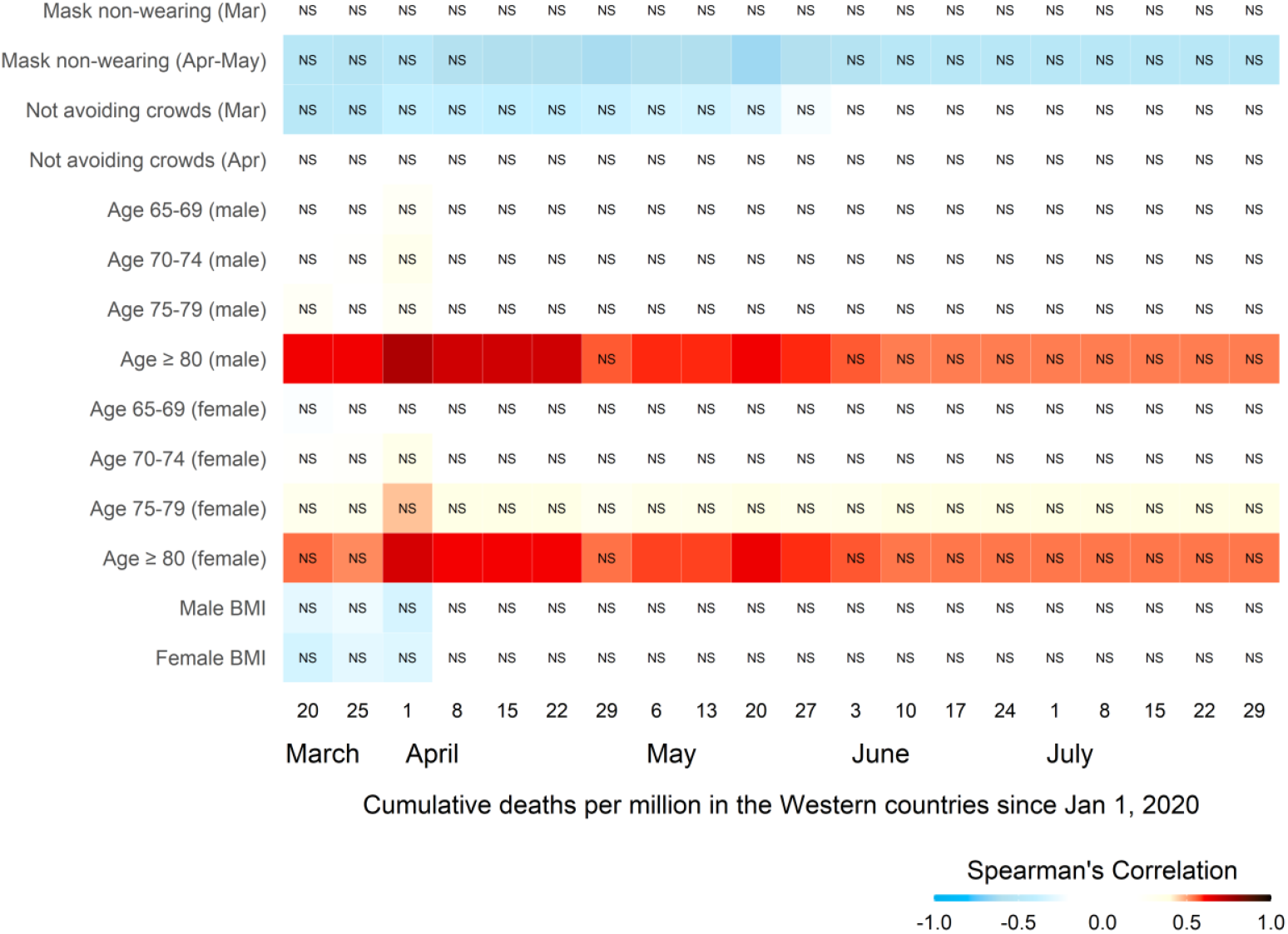
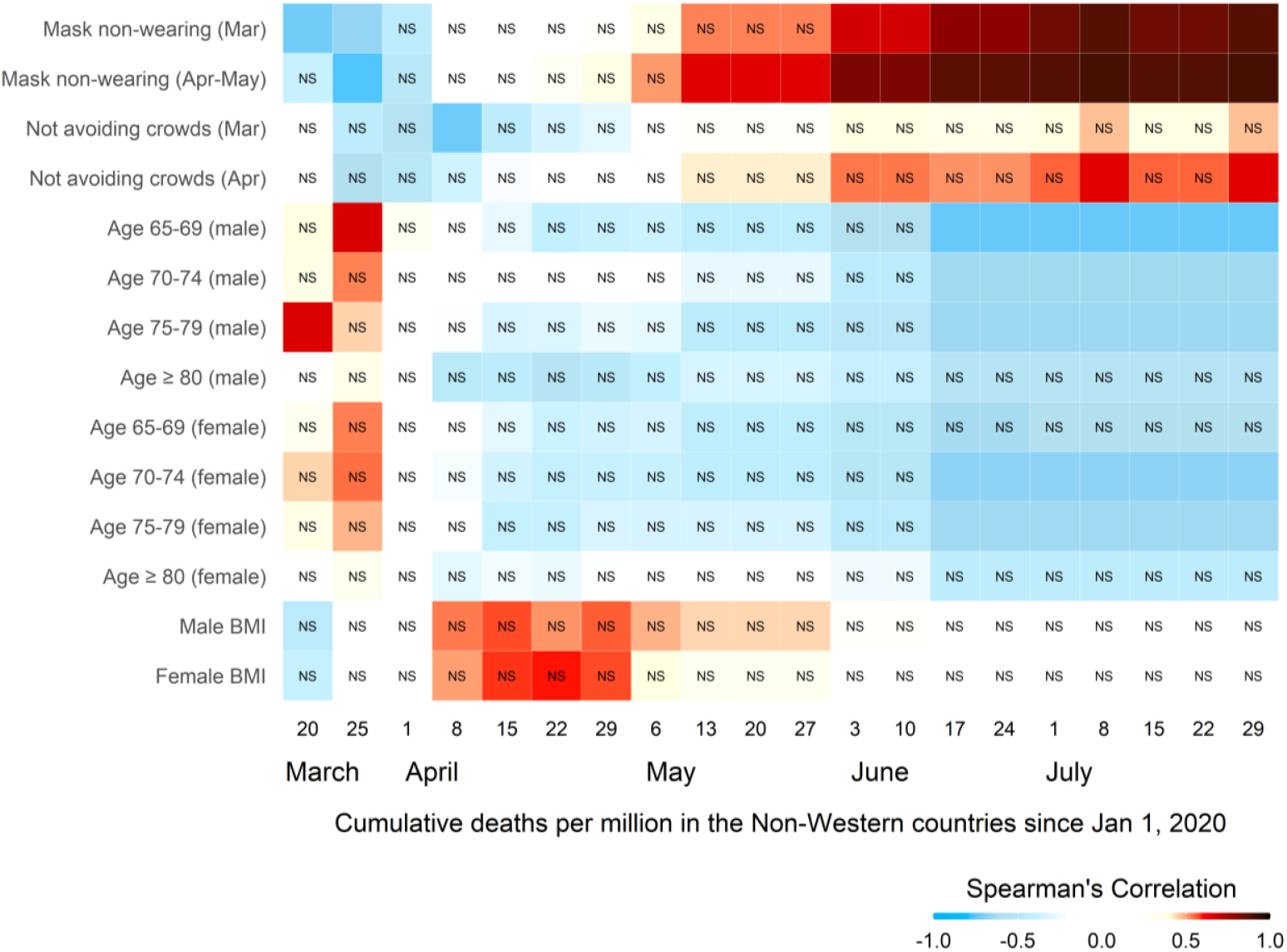
Spearman’s correlation with the cumulative number of deaths in (a) Western countries and (b) non-Western countries. Mask-wearing rates in March and April–May were calculated from the survey responses from March 9–18, 2020, and April 26–May 1, 2020, respectively. Age-related predictors indicate the percentage of the population in a specified range. BMI, body mass index; NS, not significant (*P* > 0.05). Western countries include the United Kingdom, France, Italy, USA, Spain, Germany, Canada, Sweden, Norway, Finland, Denmark, and Australia, whereas non-Western countries include Mexico, Malaysia, China, Saudi Arabia, India, Indonesia, the Philippines, Japan, Singapore, and Thailand.

In non-Western countries (Fig. 2B, Data S1), the mask non-wearing rates of March and April– May showed a significant negative correlation in March when the effects of wearing a mask were not yet expected to show. This reverse correlation may represent reverse causation caused by a fear of the disease. Later, they showed a time-dependent increase, leading to high correlations. Not avoiding crowds in non-Western countries tended to negatively correlate in the early stage of the pandemic, possibly representing reverse causation caused by fear. Later, they showed a time-dependent increase. Age-related variables in the non-Western countries tended to be positively correlated only in March. Interestingly, age-related variables in the non-Western countries showed a time-dependent decrease, leading to significant negative correlations in both sexes.

Next, we created scatter plots to visualize the relationship between several variables showing high Spearman’s correlations and the log-transformed cumulative number of deaths per million on April 1, May 13, and July 29 (Fig. 3A) and the log-transformed weekly number of deaths per million of March 30–April 5, May 11–May 17, and July 27–August 2 (Fig. 3B). April 1 and March 30–April 5 were chosen as the earliest times for mask-wearing to take effect. July 29 and July 27–August 2 were chosen as the last period for the observation. May 13 was chosen as the date with the highest Spearman’s correlation coefficient (0.739) for mask non-wearing (March), and May 11–May 17 was chosen as the period close to May 13. They showed monotonic correlations of the log-transformed value with most variables with clear separations between the Western and non-Western countries (Fig. 3C).

**Fig. 3A, 3B, and 3C.**
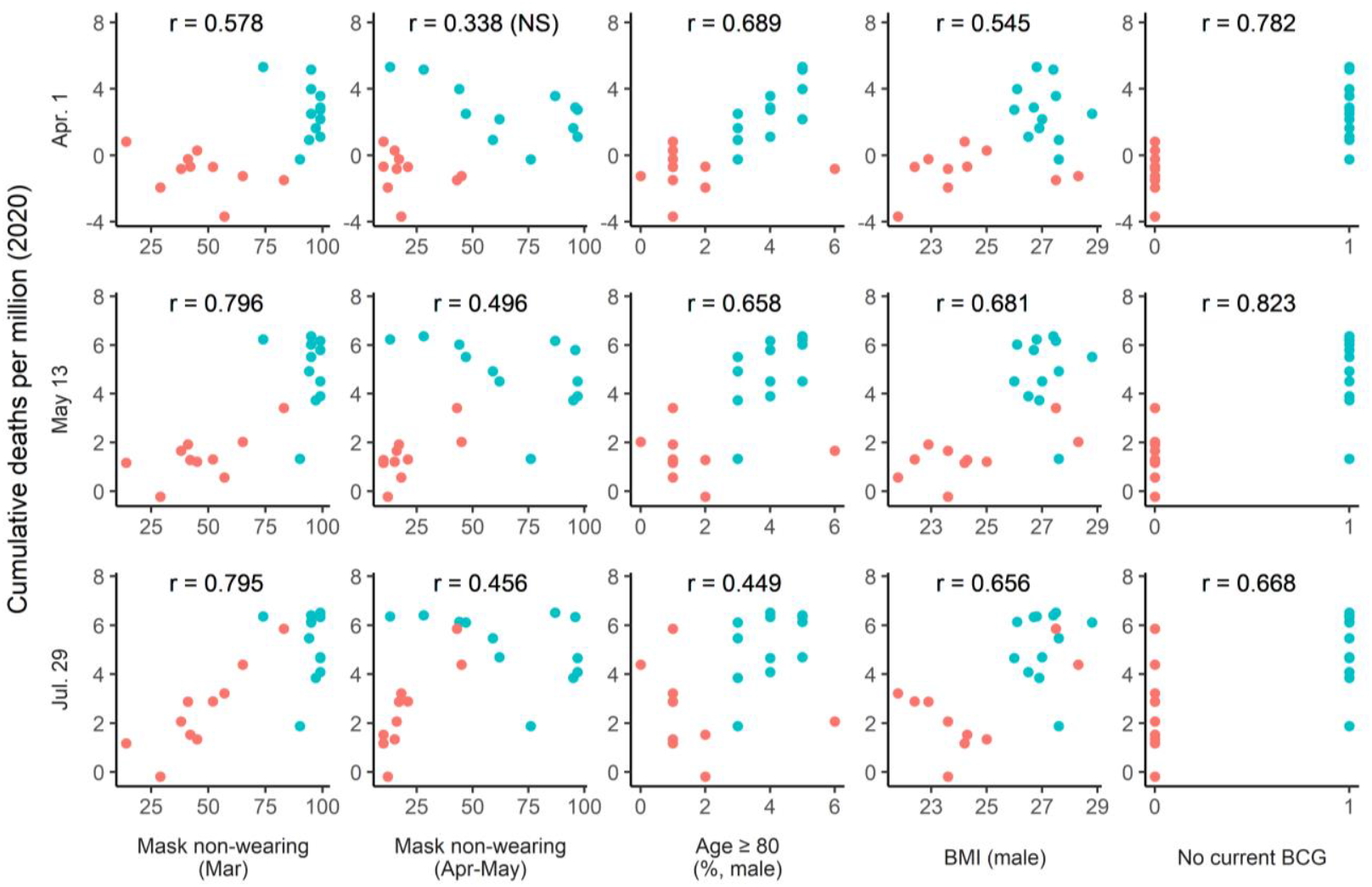
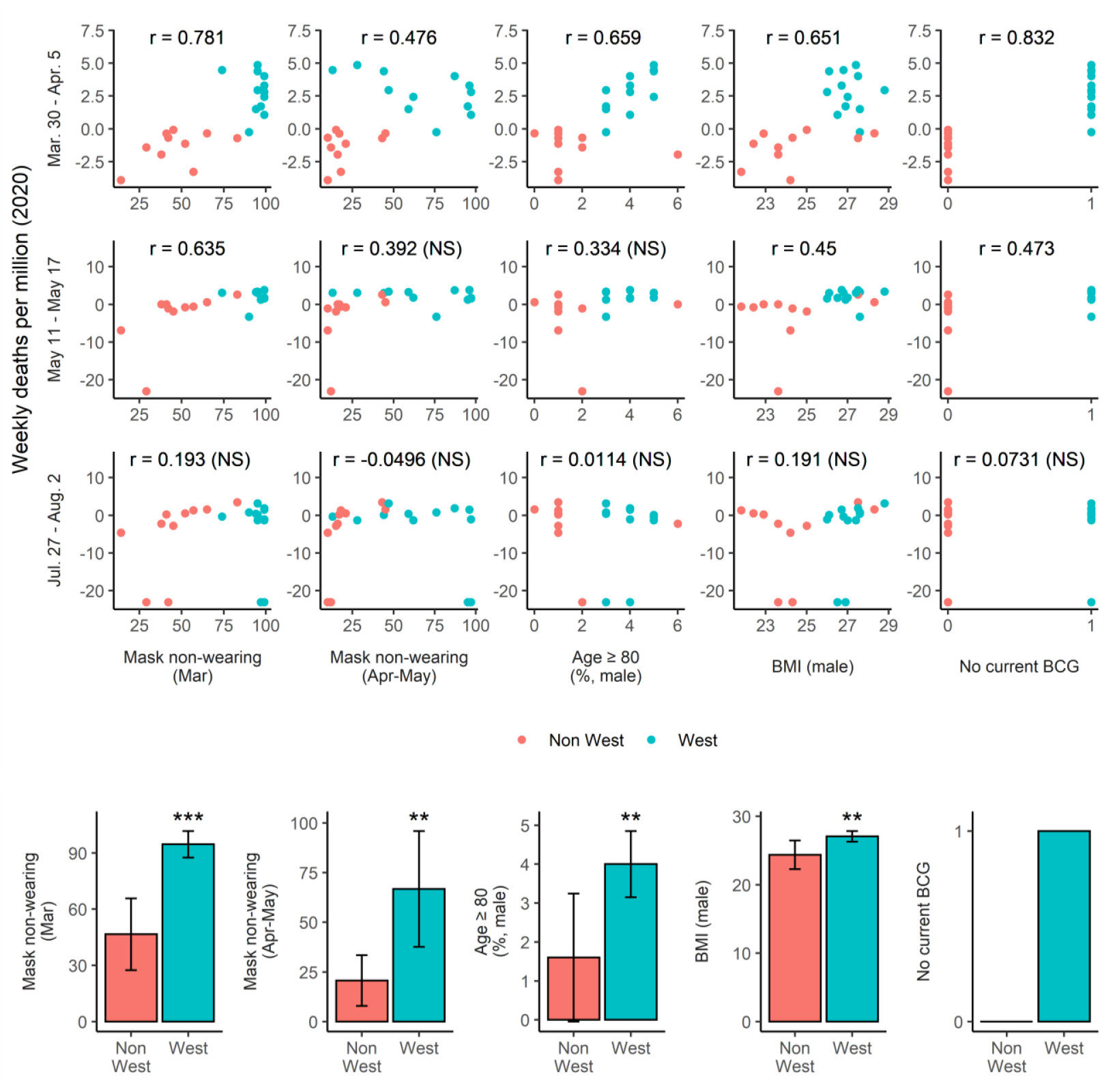
Scatter plots visualizing the correlations between (A) the predictors and the log-transformed cumulative number of deaths per million and (B) the log-transformed weekly number of deaths per million. NS, not significant (*P* > 0.05). (C) Western countries have higher mask non-wearing rates, a percentage of individuals ≥80 years, and male BMI than non-Western countries, and 100% for no current BCG. \*\**P* < 0.01, \*\*\**P* < 0.001 in Wilcoxon rank-sum test. Western countries include the United Kingdom, France, Italy, USA, Spain, Germany, Canada, Sweden, Norway, Finland, Denmark, and Australia, whereas non-Western countries include Mexico, Malaysia, China, Saudi Arabia, India, Indonesia, the Philippines, Japan, Singapore, and Thailand.

It is noted that we found a significant correlation between the variables, indicating the possibility of confounding between the variables. (Fig. S1).

We subsequently attempted to explain the log-transformed cumulative number of deaths per million on May 13 (CUMU) and the log-transformed weekly number of deaths per million from March 30–April 5 (WEEK), using the multiple linear regression approach because they were highly correlated among the cumulative and weekly groups, respectively.

We chose the March mask non-wearing rate (MASK), current BCG vaccination policy (BCGP), which represents Western countries in this study, males over 80 years (MAGE), and the BMI of males (MBMI) as explanatory variables (Table S1, Fig. 4). Regressions for other dates and weeks are shown in Table S2.

**Fig. 4.**
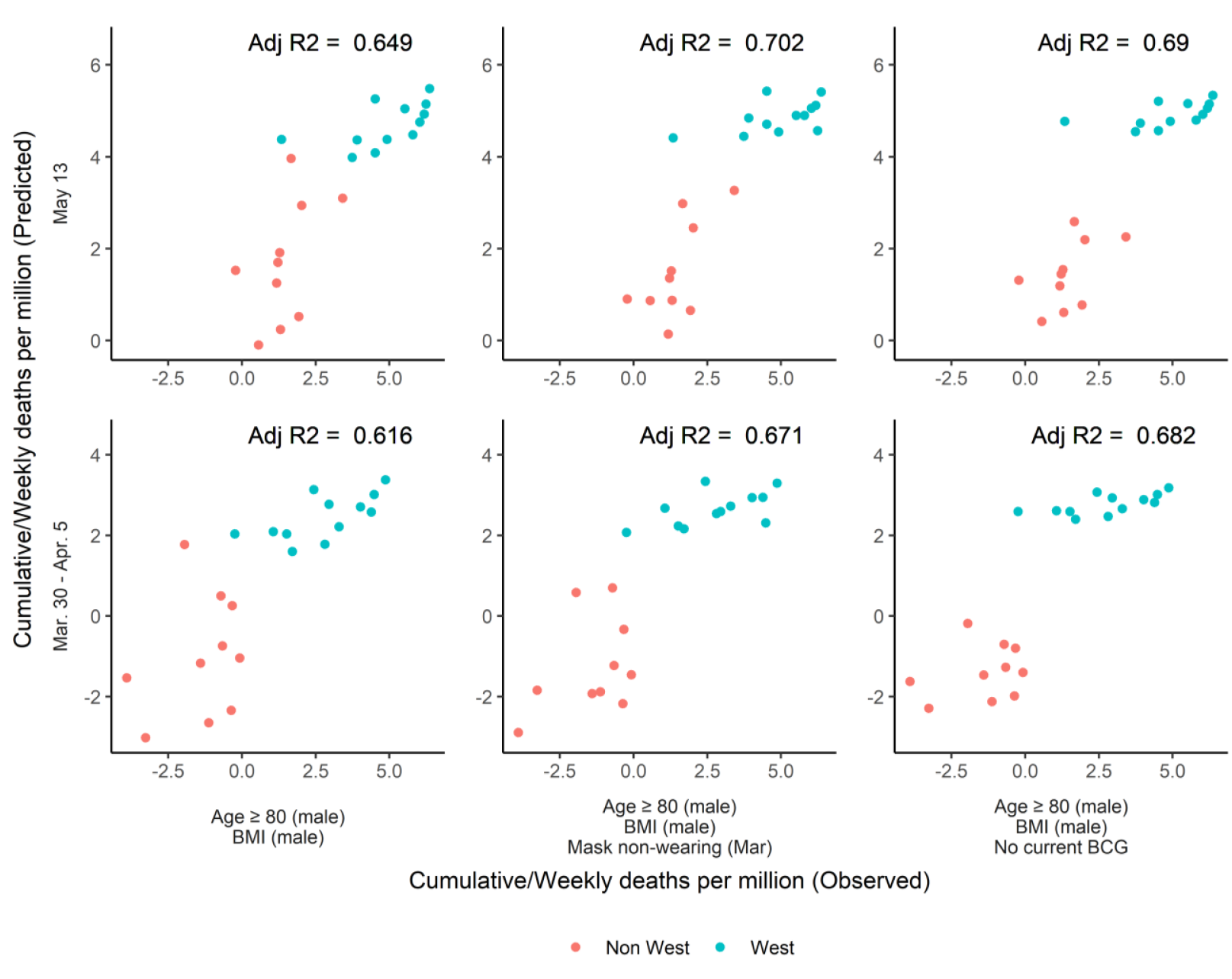
Multiple linear regression. The predicted (left: male age ≥80 + male BMI; middle: male age ≥80 + male BMI + Mask non-wearing in March; right: male age ≥80 + male BMI + no current BCG); and observed cumulative (upper) and weekly (lower) number of deaths per million were highly correlated.

MAGE was significantly associated when BCGP was not included in the model. MBMI was significantly associated when neither MASK nor BCGP was included in the model. MASK was significantly (almost significant in Models 9 and 20) associated with BCGP and was not included in the model. BCGP was significant (almost significant in Model 5), except when all three other explanatory variables were included in the model.

MAGE and MBMI contributed to 64.9% of CUMU (Fig. 4, upper left) and 61.6% of WEEK (Fig. 4, lower left), with both explanatory variables being significant. When MASK was added to the above models, the contribution rate increased to 70.2% for CUMU (Fig. 4, upper middle) and 67.1% for WEEK (Fig. 4, lower middle). Similarly, when BCGP was added to the models, the contribution rate increased to 69.0% for CUMU (Fig. 3, upper right) and 68.2% for WEEK (Fig. 4, lower right). Based on these results, it can be said that MASK and/or BCGP could be an additional explanatory factor.

Each coefficient of determination (R2) is described as VIF=1/(1-R2). MASK was 64.7% (R2=0.647, Models 9 and 20), as explained by MAGE and MBMI. BCGP was 72.9%, as explained by MAGE and MBMI. MASK and BCGP explained 76.5% of each other. BCGP was 85.6%, explained by the MASK, MAGE, and MBMI, while MASK was 81.2%, explained by BCGP, MAGE, and MBMI. The adjusted R2 of Model 11 (0.692) did not improve when compared to Model 9 (0.702) or Model 10 (0.69). The adjusted R2 of Model 22 (0.674) did not improve when compared to Model 20 (0.671) or Model 21 (0.682). These results indicate that most of the effects of MASK on mortality are explained by the other three variables, and the same can be said for BCGP. Thus, if we consider MASK and BCGP as candidates for additional explanatory factors for MAGE and MBMI, it is unlikely that both are meaningful at the same time; therefore, one of them may not be an explanatory factor. If BCGP is not an explanatory factor, then the hypothesis shown in Fig. S2 may be considered. The following are some findings that may indicate that BCGP is not an explanatory factor. In Western countries that do not currently offer BCG vaccination, a positive correlation between age ≥80 years in both sexes and cumulative deaths was observed (Fig. 2A). In non-Western countries that currently offer BCG vaccinations, a positive correlation between mask-wearing rates of March and April–May and cumulative deaths was observed (Fig. 2B). Note that in Fig. 1B, correlations of age-related variables and no current BCG diminished in late May, and the mask non-wearing rate (March and April–May) diminished in late June, but BMIs tended to last until the end of the observation period. If it is assumed that the mask-wearing rate at a point contributes to transmission at that point in time, the resulting deaths may be stronger at a subsequent time. Factors such as age, obesity, and BCG status, which were present and unchanged during the observation period, should be constantly associated with mortality over the period of observation, unless people with these characteristics change their behavior, such as increased preventive behavior. People in the higher BMI group may not have changed their behavior over the observation period; therefore, the correlation tended to last until the end of the observation period. In Japan, older people have been reported to have taken stronger preventive behavior than younger people^16^; this may also be true in other countries, especially in non-Western countries^15^, and could be the reason why correlations with age-related variables diminished in later periods (Fig. 1B) and why negative correlations were found among non-Western countries in later periods (Fig. 2B). Assuming that childhood BCG vaccination affects lifelong immunity and resistance to COVID-19, we could not find a convincing explanation for a sharp decrease in the correlation with weekly deaths after May. This diminishing pattern is similar to the patterns of age-related factors and mask non-wearing rates, which may indicate that they may be confounding factors of BCG vaccination policy. This is consistent with a report questioning the effect of BCG vaccination on the grounds that there was a correlation between BCG vaccination and COVID-19 deaths in April, but not in August *(17)*. Although our variable for BCG is the division only of the current childhood vaccination policy and did not consider more detailed information, such as the timing of the establishment or discontinuation of vaccination in each country, its correlation with COVID-19 deaths by country was originally focused on because it correlated with division only of current status ^11^.

We observed considerable differences in face mask-wearing rates between Western and non-Western countries. We speculate that each country’s policy regarding wearing face masks cannot explain this clear difference because, for example, face masks were never mandated in Japan, despite the high face mask-wearing rate^18^. We speculate that cultural factors could be one of the reasons for the difference in mask-wearing rate, as Jack et al.^19^ described it as follows, “whereas Western Caucasian internal representations predominantly featured the eyebrows and mouth, East Asian internal representations showed a preference for expressive information in the eye region.” This tendency could explain why it is considered rude to wear sunglasses among East Asians and suspicious to wear face masks in Western countries^20^.

A major limitation of this study is that we were able to obtain synchronized mask-wearing rates from only 22 countries. More synchronized and detailed data on mask-wearing rates, such as age and sex, should be collected for a more detailed analysis. In addition, the data in this study were ecological and, therefore, subject to potential ecological fallacies. Data that repeatedly measure the rate of mask use within each country over time, including more information on confounding factors or the introduction of interventions, are needed to address the causality.

Taken together, the present study demonstrated the close association between face mask-wearing rates, especially in the early phase of the pandemic (mid-March), current BCG vaccination policy, age ≥80 years, BMI, and the number of deaths caused by COVID-19, with clear differences between Western and non-Western countries. Confounding was inferred from the variables. We have shown that face mask-wearing in March, age ≥80 years of men, and BMI of males, together explained 70.2% of the cumulative number of COVID-19 deaths per million on May 13 and 85.6% of the current BCG vaccination policy. We have shown that it is unlikely that both face mask-wearing in March and the BCG vaccination policy are meaningful at the same time and proposed the hypothesis shown in Fig. S2. Although there may be other unknown confounders with clear separations between Western and non-Western countries, and the study does not prove a causal relationship between observed variables, these findings provide clues to unravel the mystery of differences in mortality by country. It has implications for the introduction of mandatory face mask use as a precautionary principle, especially in the early phase of a pandemic, based on the assumption of a causal relationship.

## Data Availability

All data was collected from publicly available secondary sources.
% of people in each country who say they are: Wearing a face mask when in an public spaces as face mask wearing rate in mid-March(3/9-18) 2020 and late April to early May(4/26-5/1) 2020 of countries listed below were obtained from:
Smith M. International COVID-19 Tracker Update: 2 May. 2 May 2020, yougov.co.uk/topics/international/articles-reports/2020/05/01/international-covid-19-tracker-update-2-may
Smith M. International COVID-19 Tracker Update: 18 May. 18 May 2020, yougov.co.uk/topics/international/articles-reports/2020/05/18/international-covid-19-tracker-update-18-may
Total COVID death per million of countries listed below except for Taiwan were obtained from:
Ritchie, Hannah. Coronavirus Source Data. 16 June 2020, ourworldindata.org/coronavirus-source-data
Data of BMIs of countries listed below except for Taiwan were obtained from:
Global Status Report on Noncommunicable Diseases 2014. 5 Oct. 2015, www.who.int/nmh/publications/ncd-status-report-2014/en/.
Population% by age of countries listed below except for Taiwan were obtained from:
Kose, Ayhan, et al. World Bank Open Data. Data, 15 June 2020, data.worldbank.org/.

https://github.com/kusami1/mask

## Materials and Methods

All data were collected from publicly available secondary sources. Analyses in the present study included 12 Western countries (UK, France, Italy, USA, Spain, Germany, Canada, Sweden, Norway, Finland, Denmark, and Australia) and 10 non-Western countries (Mexico, Malaysia, China, Saudi Arabia, India, Indonesia, Philippines, Japan, Singapore, and Thailand). These countries were chosen because of the availability of synchronized March mask-wearing data. The face mask-wearing rates in March (March 9 to 18) 2020 and from late April to early May (April 26 to May 1) 2020 across countries were derived from the item “percentage of people in each country who answered that they are wearing a face mask when in public spaces.” The rate of avoiding crowds in March (March 13 to 19) 2020 and from late April to early May (April 26 to May 1) 2020 across countries were derived from the item “% of people in each market who say they are: Avoiding crowded public spaces” from https://yougov.co.uk/topics/international/articles-reports/2020/03/17/personal-measures-taken-avoid-covid-19. The rate for Australia from late April to early May was estimated to be 78% from the data of March 29 (80%) and May 22 (76%). This YouGov database has collaborated with the Institute of Global Health Innovation at Imperial College London and summarizes interviews conducted with nationally representative samples (150–2000/week depending on the country). Total COVID-19 deaths per million were obtained from https://ourworldindata.org/coronavirus-source-data. The BMI data were obtained from the Global Status Report on Non-communicable Diseases 2014^1^ (October 5, 2015; https://www.who.int/nmh/publications/ncd-status-report-2014/en/). Population percent by age data was obtained from https://data.worldbank.org on June 15, 2020. BCG vaccination policies were obtained from http://www.bcgatlas.org/index.php.

### Data Analysis

Statistical analyses were conducted using R version 4.0.0. The R packages used in this study include ggplot2, car, ggcorrplot, lemon, ggpubr, Hmisc, rstatix, and lubridate. Spearman’s correlations were calculated using the “rcorr” functions. Linear regression models were built using the “lm” function.

The data used for analysis is available at https://github.com/kusami1/mask

## Data and materials availability

All data and codes are available in the main text or supplementary materials.

## Methods References

## Acknowledgments

The author would like to express sincere thanks to Dr. Gen Kaneko, Assistant Professor of Biology, School of Arts and Sciences, University of Houston-Victoria and Mr. Junya Iwai (journalist) for their help with statistical assistance, data collection, and constructive discussions.

## Author contributions

**Daisuke Miyazawa**: conceptualization, data curation, project administration, resources, writing, methodology, formal analysis, validation, **and visualization**.

## Competing interests

None

## Supplementary Materials

Figs. S1 to S2

Tables S1 to S2

Data S1

**This PDF file includes**

Figs. S1 to S2

Tables S1 to S2

**Other Supplementary Materials for this manuscript include the following**

Data S1 [Spearman’s correlations]

**Fig. S1.**
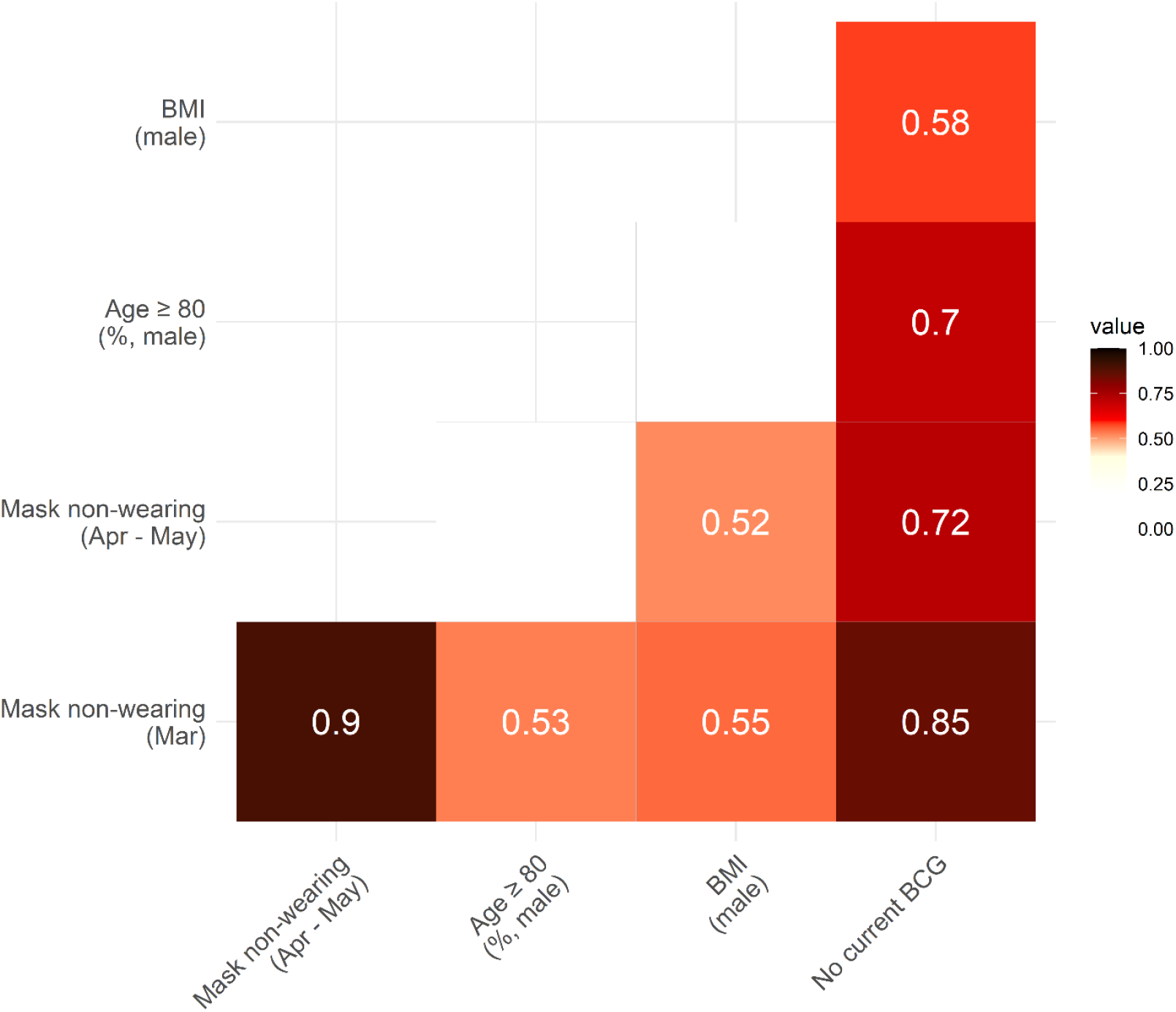
Spearman’s correlation matrix between predictors. Only significant correlations are shown with rho values (*P* < 0.05).

**Fig. S2.**
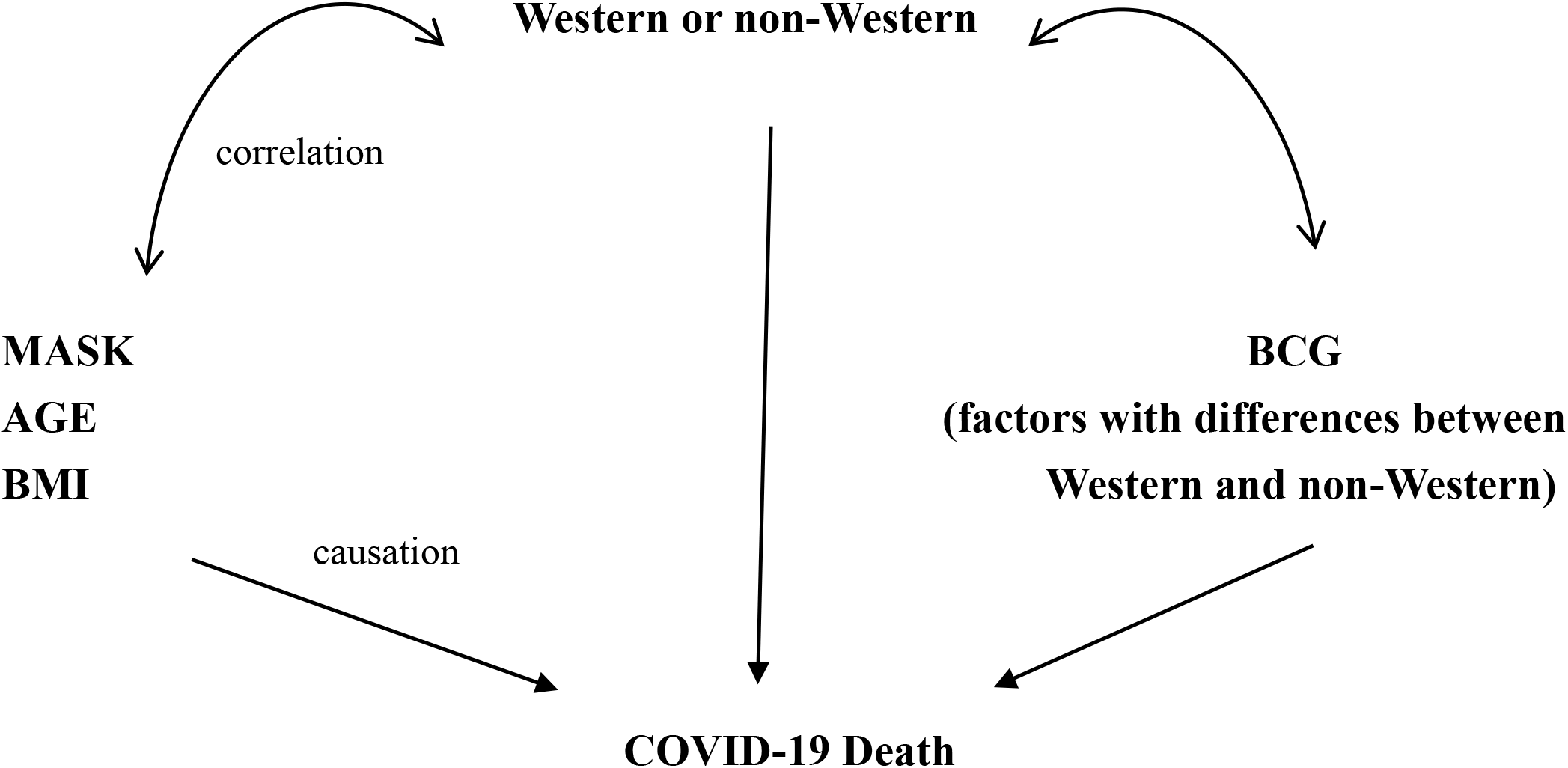
Hypothesis that BCG policy is not an explanatory factor, but wearing face masks, old age, and obesity are.

**Table S1.**
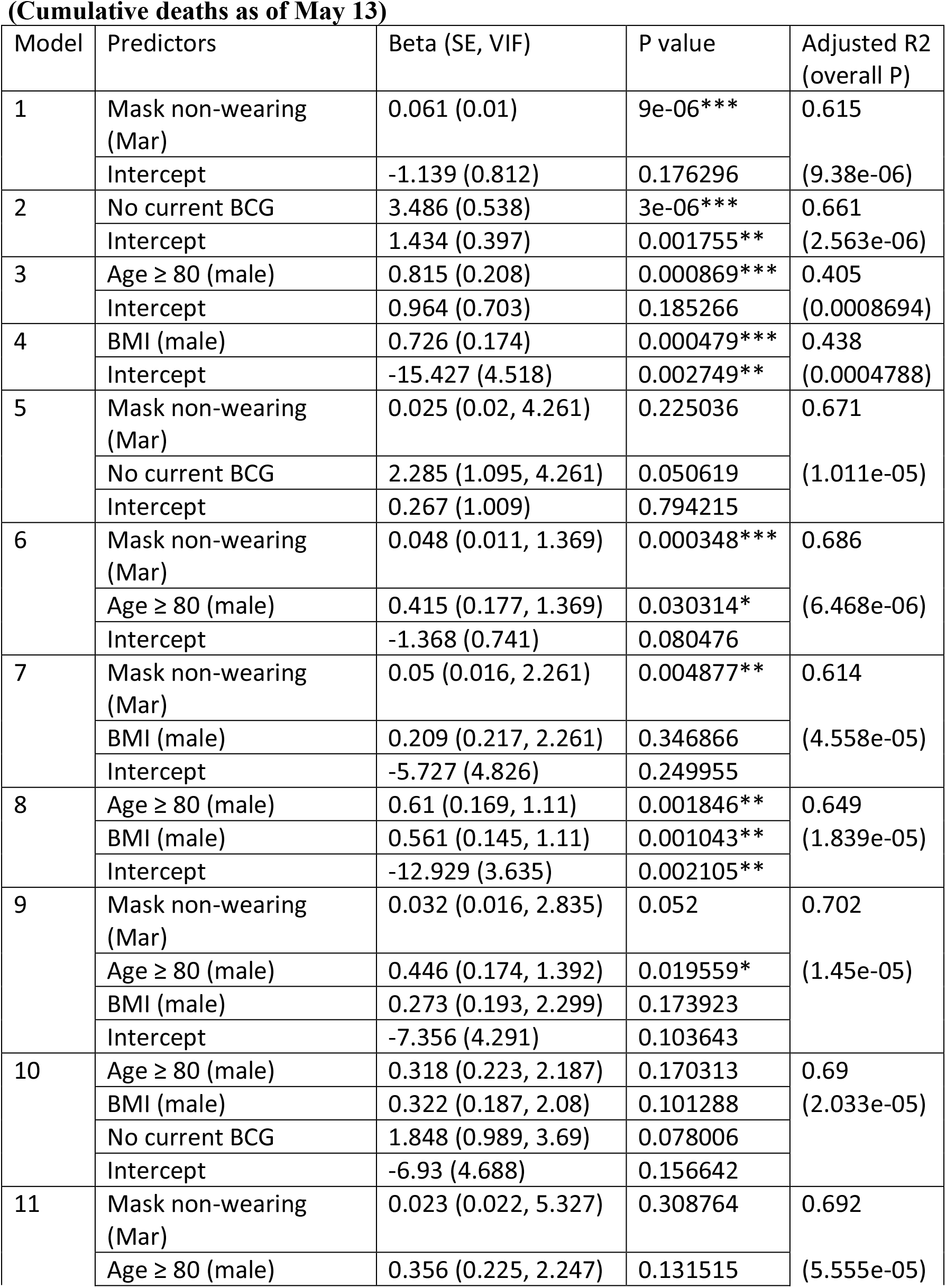
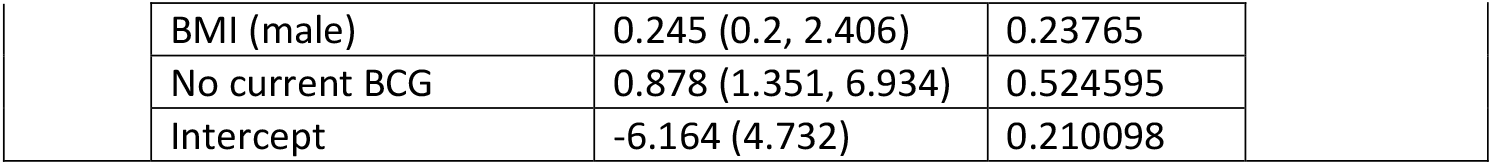
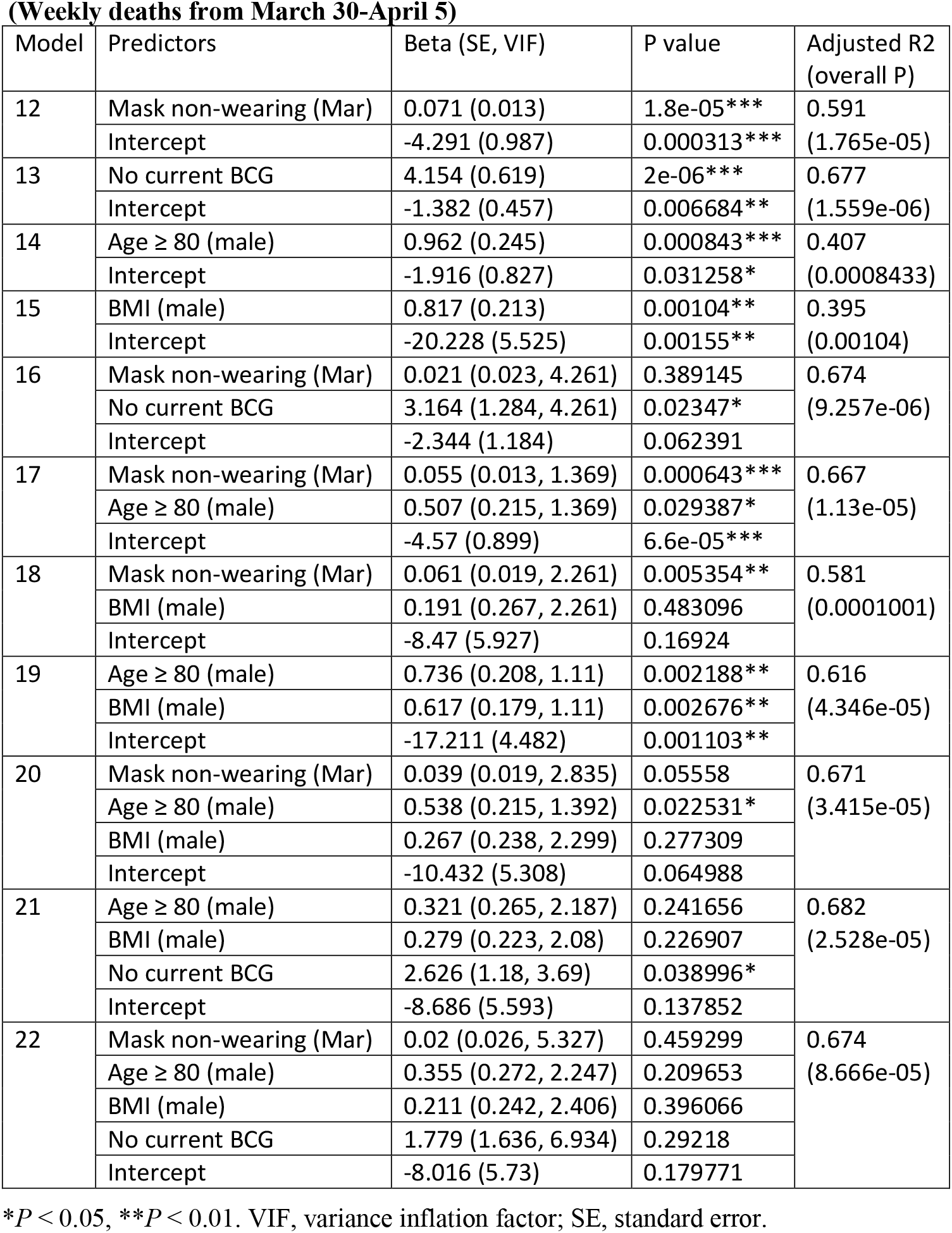
Linear regression

**Table S2.**
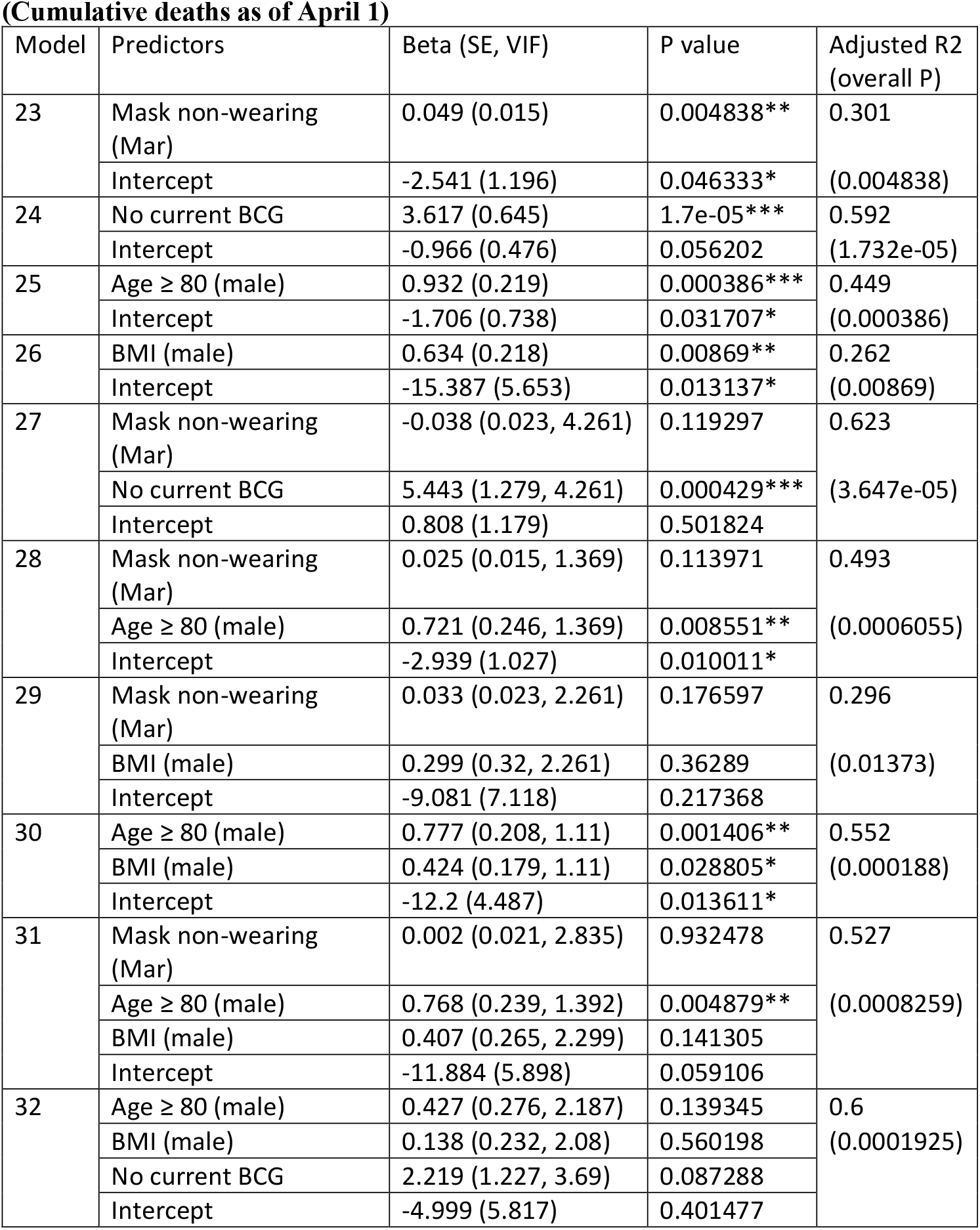
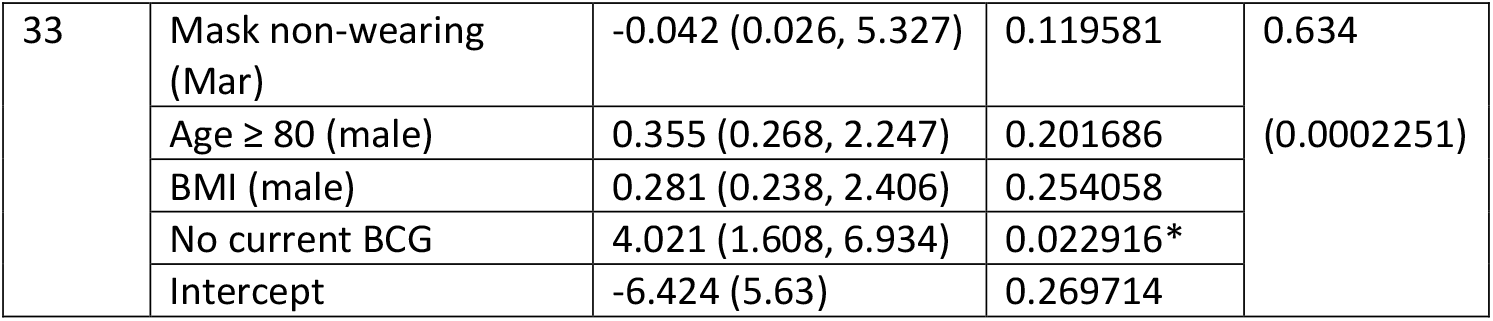
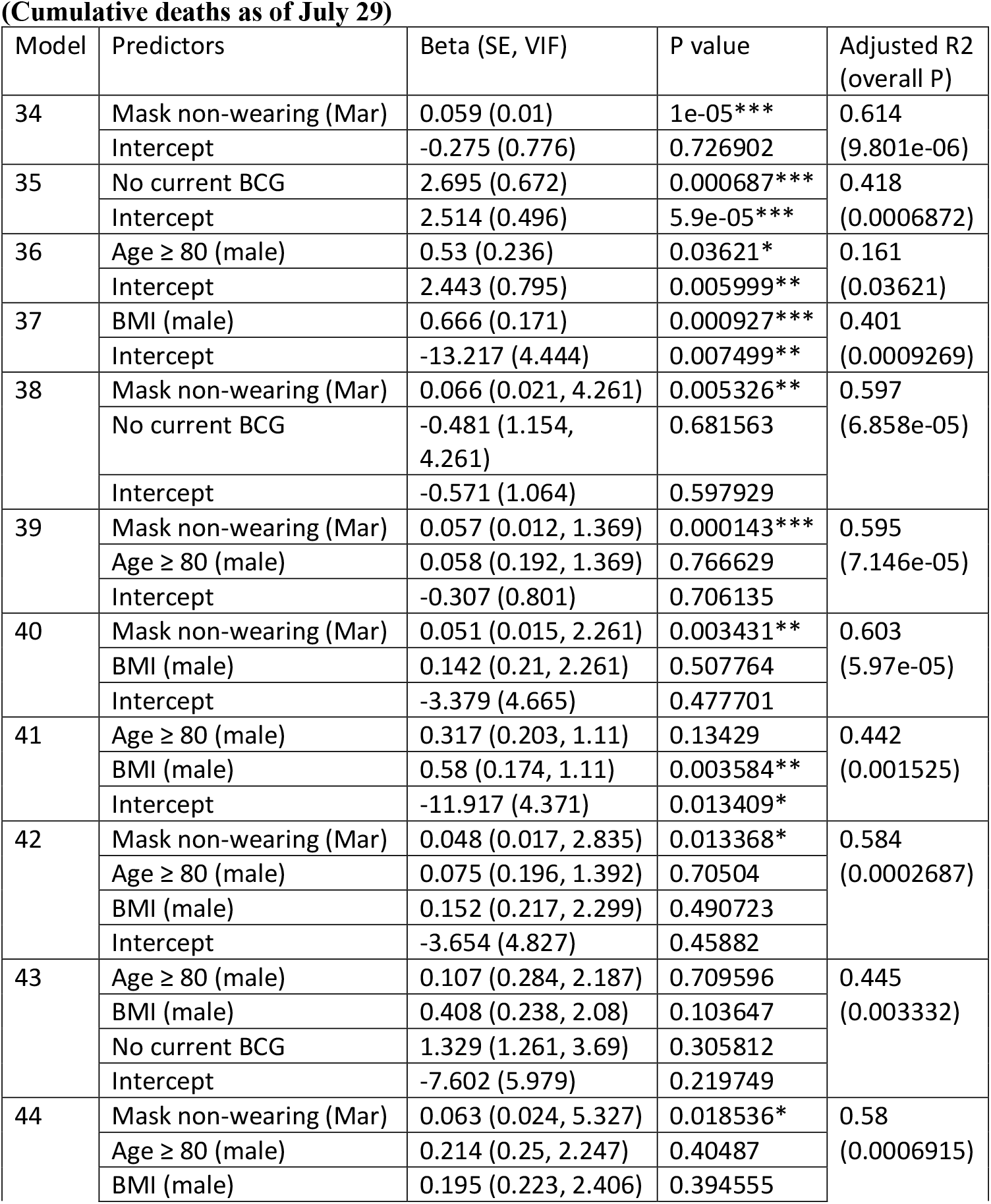
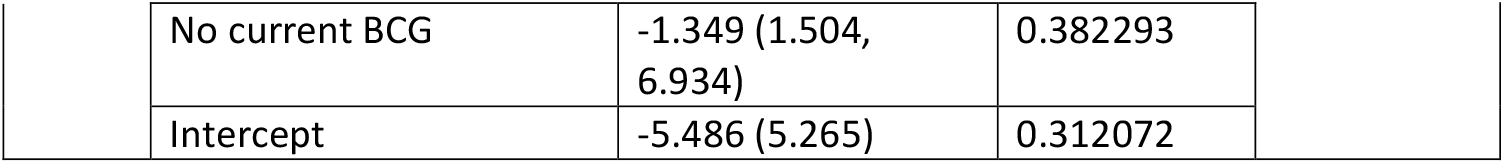
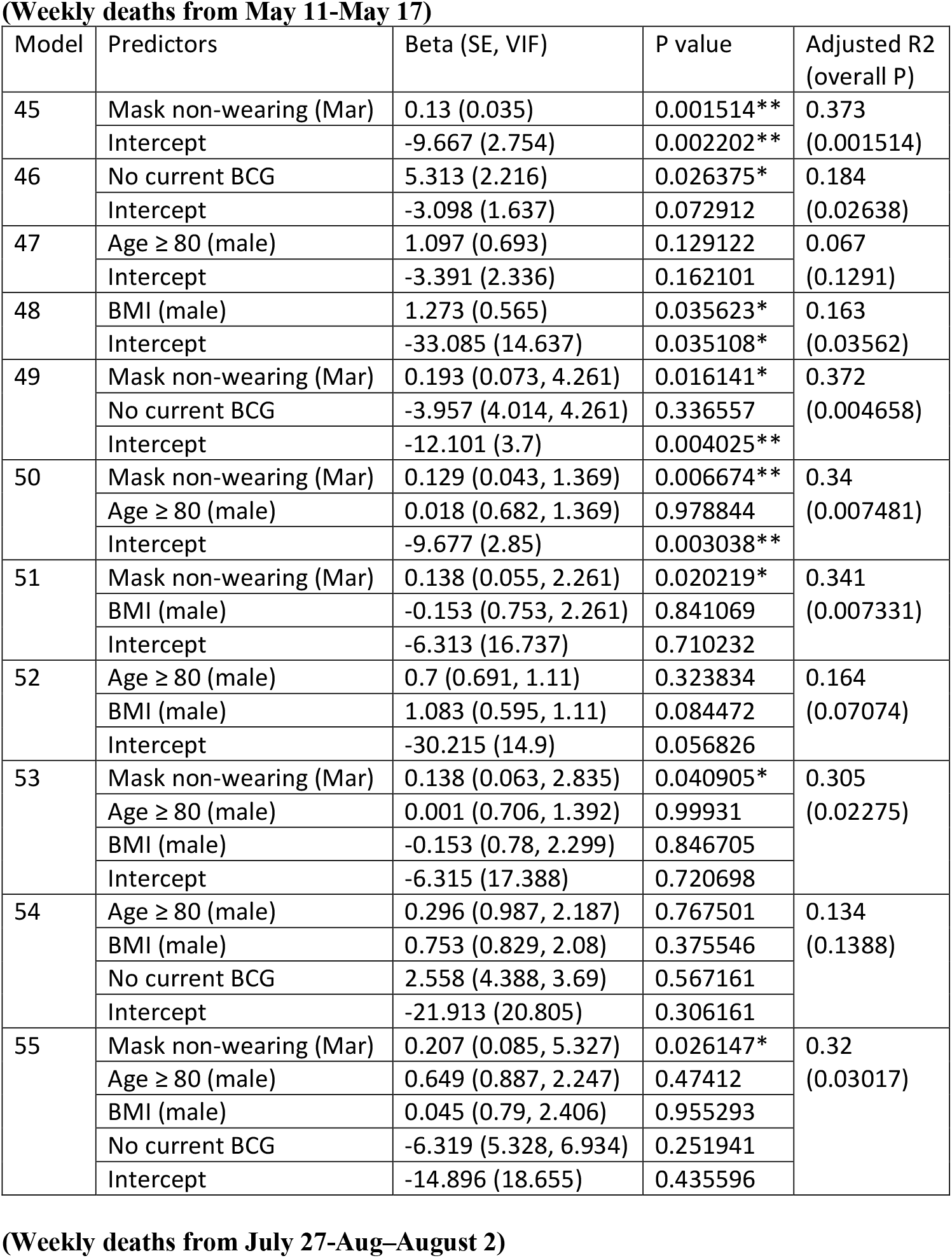
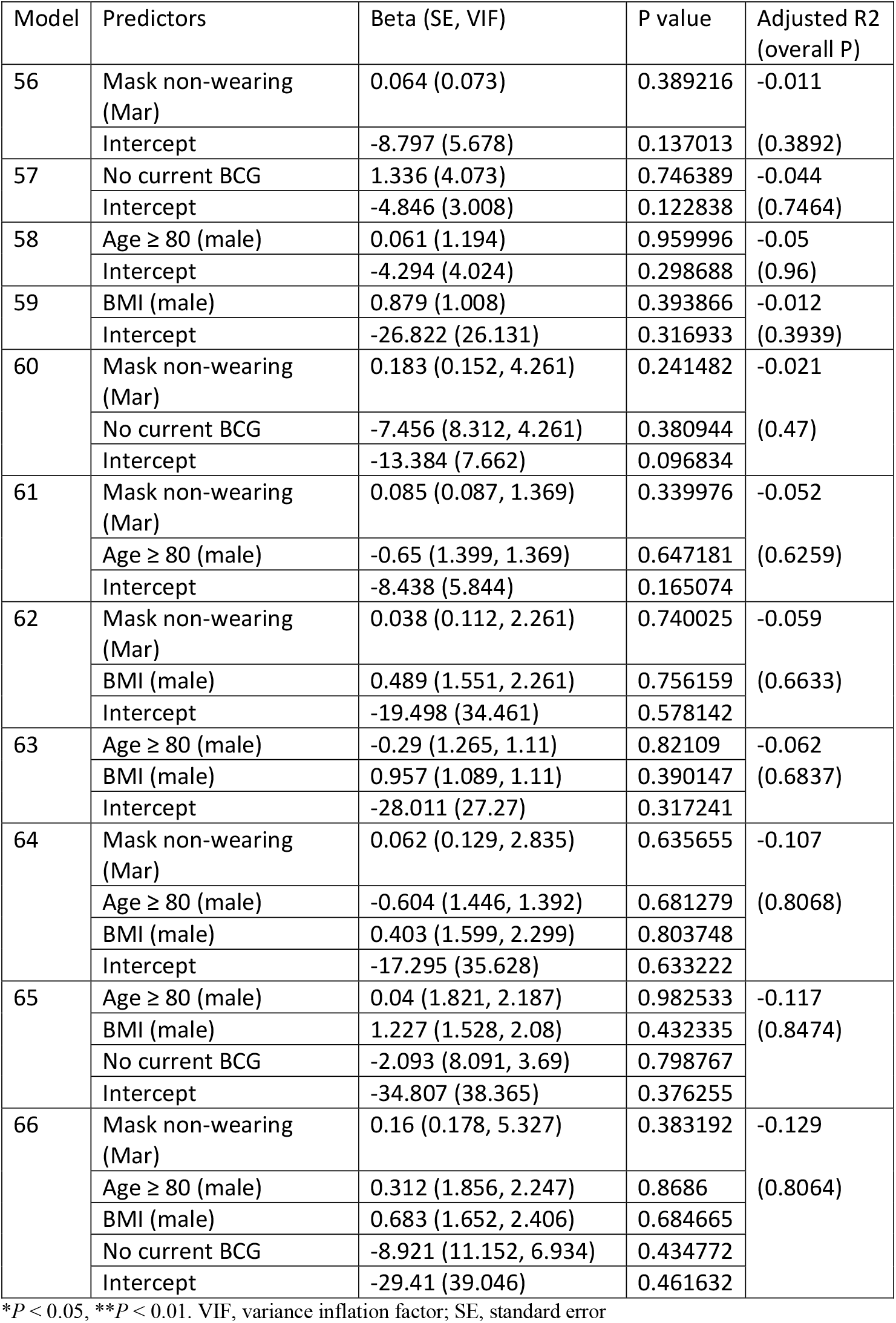
Linear regressions

**Data S1. (separate file)**

Spearman’s correlations

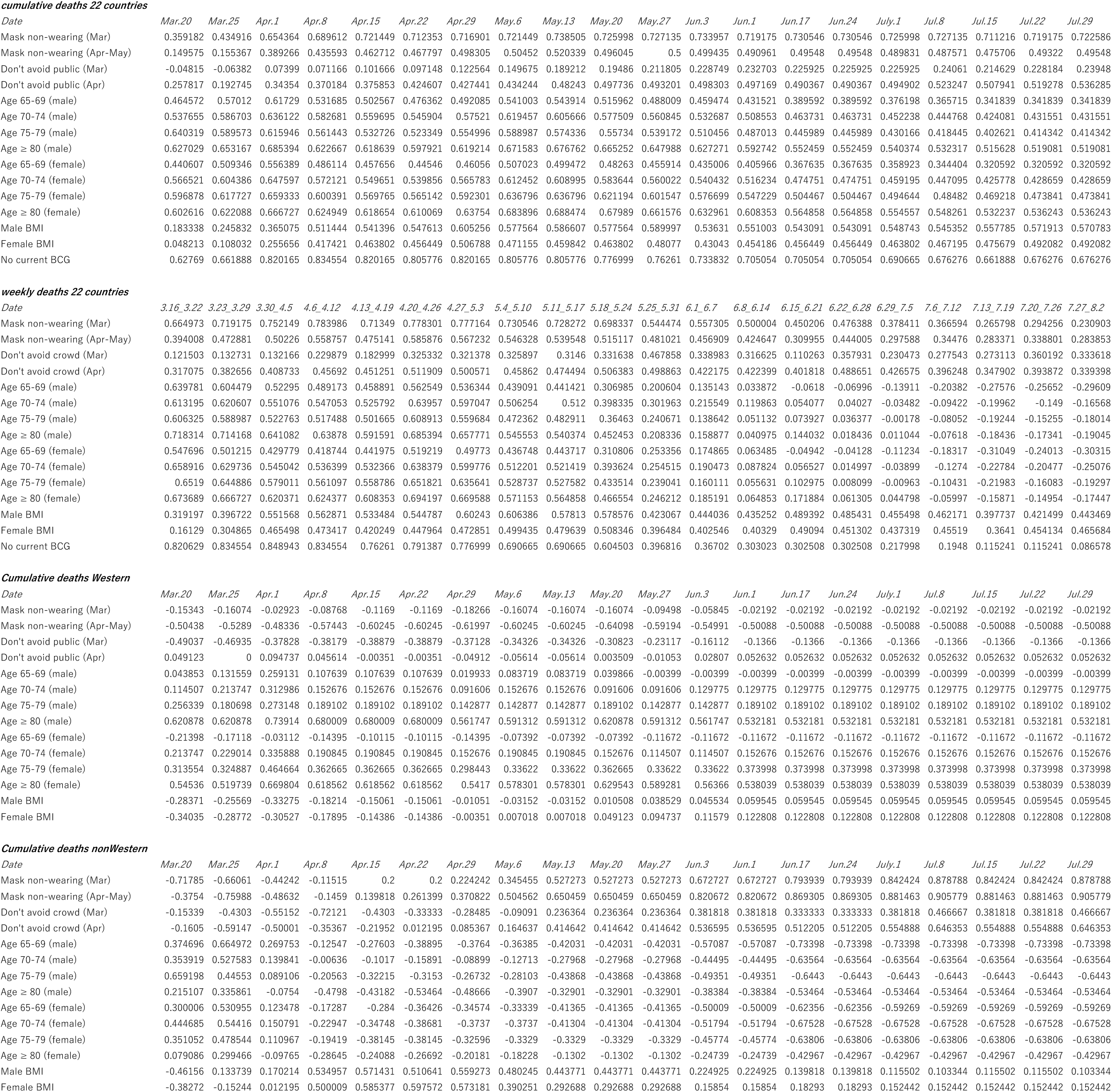

## Notes

### Competing Interest Statement

The authors have declared no competing interest.

### Funding Statement

The author(s) received no specific funding for this work.

### Author Declarations

All information used for this study has been published. There is therefore no ethical issue related to this study.

### Summary of Updates

We also proposed the hypothesis that these variables may be confounders of other suspected factors with large differences between Western and non-Western countries, such as the BCG vaccination policy.

## References

1. Leung, N. Y., Bulterys, M. A., Bulterys, P. L. Predictors of COVID-19 incidence, mortality, and epidemic growth rate at the country level. Preprint at https://doi.org/10.1101/2020.05.15.20101097 (2020).

2. Squalli, J. Evaluating the determinants of COVID-19 mortality: A cross-country study. Preprint at https://doi.org/10.1101/2020.05.12.20099093 (2020).

3. Knittel, C.R. Ozaltu, B. What does and does not correlate with COVID-19 death rates. Preprint at https://doi.org/10.1101/2020.06.09.20126805 (2020).

4. Leffler, C. T. et al., A. Association of Country-wide Coronavirus Mortality with Demographics, Testing, Lockdowns, and Public Wearing of Masks. The American journal of tropical medicine and hygiene, 103(6), 2400–2411. https://doi.org/10.4269/ajtmh.20-1015 (2020).

5. Chu, D. K. et al., Physical distancing, face masks, and eye protection to prevent person-to-person transmission of SARS-CoV-2 and COVID-19: a systematic review and meta-analysis. Lancet 395, (10242), 1973–1987, (2020).

6. Centers for Disease Control and Prevention, Scientific Brief: Community Use of Cloth Masks to Control the Spread of SARS-CoV-2, https://www.cdc.gov/coronavirus/2019-ncov/more/masking-science-sars-cov2.html (2020).

7. Miyazawa, D., Why obesity, hypertension, diabetes, and ethnicities are common risk factors for COVID-19 and H1N1 influenza infections. J. Med. Virol. https://doi.org/10.22541/au.159181185.59632853 (2020).

8. Mueller, A. L., McNamara, M. S., Sinclair, D. A., Why does COVID-19 disproportionately 10 affect older people?. https://doi.org/10.18632/aging.103344 (2020).

9. Petrilli, C. M. et al., Factors associated with hospital admission and critical illness among 5279 people with coronavirus disease 2019 in New York City: a prospective cohort study. BMJ 369. https://doi.org/10.1136/bmj.m1966 (2020).

10. Docherty, A. B. et al., Features of 20?133 UK patients in hospital with COVID-19 using the ISARIC WHO clinical characterization protocol: a prospective observational cohort study. BMJ 369, https://doi.org/10.1136/bmj.m1985 (2020).

11. Peduzzi, P., Concato, J., Feinstein, A. R., and Holford, T. R. Importance of events per independent variable in the proportional hazards regression analysis. II. Accuracy and precision of regression estimates. J Clin Epidemiol 48, 1503–1510, https://doi.org/10.1016/0895-4356(95)00048-8 (1995).

12. Miller, A. et al. Correlation between universal BCG vaccination policy and reduced mortality for COVID-19. Preprint at https://doi.org/10.1101/2020.03.24.20042937 (2020).

13. Montopoli, M. et al., Androgen-deprivation therapies for prostate cancer and risk of infection by SARS-CoV-2: a population-based study (N= 4532). Ann. Oncol. 31, https://doi.org/10.1016/j.annonc.2020.04.479 (2020).

14. Grandi, G., Facchinetti, F., Bitzer, J., The gendered impact of coronavirus disease (COVID-19): do estrogens play a role? Eur. J. Contracept. Reprod. Health Care 25, 233–234, https://doi.org/10.1080/13625187.2020.1766017 (2020).

15. Yildirim, M., Geçer, E., & Akgül, Ö. (2021). The impacts of vulnerability, perceived risk, and fear on preventive behaviours against COVID-19. Psychology, health & medicine, 26(1), 35–43. https://doi.org/10.1080/13548506.2020.1776891 (2021).

16. Muto, K., Yamamoto, I., Nagasu, M., Tanaka, M., Wada, K.. Japanese citizens’ behavioral changes and preparedness against COVID-19: An online survey during the early phase of the pandemic, PLoS One 15, e0234292 https://doi.org/10.1371/journal.pone.0234292 (2020).

17. Lindestam Arlehamn C.S., Sette, A., Peters, B. Lack of evidence for BCG vaccine protection from severe COVID-19. Proc Natl Acad Sci U S A. 2020;117(41):25203–25204. https://doi.org/10.1073/pnas.2016733117 (2020).

18. Miyazawa, D., Kaneko, G., Japan’s key “X-factor” for low COVID-19 mortality may be its culture of wearing face masks. Preprint at https://doi.org/10.22541/au.159373225.52467275 (2020).

19. Jack, R. E., Caldara, R., Schyns, P. G., Internal representations reveal cultural diversity in expectations of facial expressions of emotion. J. Exp. Psychol. 141, 19–25, https://doi.org/10.1037/a0023463 (2012).

20. Gesteland, R. R., Cross-cultural business behavior: A guide for global management. (Copenhagen Business School Press, Copenhagen, 2012).

